# Extracellular vesicles as biomarkers and disease mediators in lichen planus: a systematic review & meta-analysis

**DOI:** 10.64898/2026.06.29.26356896

**Authors:** Sarah Ning, Erin Suh, Hash Brown Taha

## Abstract

**Background:** Lichen Planus (LP) is a chronic inflammatory disorder that can affect the skin, hair, nails, and mucous membranes. Oral lichen planus (OLP), the most common LP subtype, is a disease of the oral mucosa, often diagnosed through clinical examination and histopathological confirmation. Extracellular vesicles (EVs) transfer proteins, lipids, and nucleic acids among cells and have become increasingly studied for their potential as minimally invasive diagnostic biomarkers and therapeutic agents in inflammatory and autoimmune diseases.

**Methods:** PUBMED and Embase were searched from inception through June 27th, 2026. Human studies investigating EV-associated miRNA or protein biomarkers in LP and its subtypes were included, with risk of bias assessed using a modified Newcastle-Ottawa Scale. Diagnostic accuracy was evaluated using receiver operating characteristic (ROC) and BRMA models when sufficient data were available.

**Results:** Ten articles met the inclusion criteria, encompassing biomarker discovery, functional, and mechanistic studies of EVs in OLP. These included studies (n = 10) comprised 298 individuals with LP (weighted mean age 50.7 years; 61.5% female) and 194 controls (weighted mean age 47.8 years; 58.5% female). OLP-specific cohorts (n = 9 studies) included 261 individuals with OLP (weighted mean age 50.7 years; 61.4% female). Although no individual EV-associated miRNAs or proteins overlapped across studies, EV-associated miRNAs demonstrated substantial heterogeneity, while EV-associated protein findings centered on pathways related to antigen presentation, inflammatory signaling, and immune activation. Several candidate biomarkers, including miR-4484, miR-34a-5p, *GJA1*, PDIA3, and Cx43, showed potential diagnostic or prognostic relevance. ROC analyses demonstrated good diagnostic utility for miR-4484 (AUC = 0.81), and the combination of GJA1 and Cx43 showed the strongest discriminatory ability (AUC = 0.892). The diagnostic accuracy meta-analysis showed good discrimination (pooled AUC = 0.89). Functional and mechanistic studies suggested that EVs may actively contribute to OLP pathogenesis through promoting epithelial injury and activating inflammatory signalling pathways.

**Conclusions:** EV-associated miRNAs and proteins are potential biomarker candidates for LP and may provide insight into the inflammatory and immune mechanisms underlying disease pathophysiology. Functional and mechanistic evidence further suggests that EVs may play an active role in disease progression. However, current evidence has limitations such as small sample sizes and methodological heterogeneity. Larger, standardized, and longitudinal studies are needed to validate candidate EV biomarkers and determine whether EVs can serve as therapeutic tools in LP.

## Introduction

Lichen Planus (LP) is a group of chronic, immune-mediated inflammatory diseases affecting stratified squamous epithelia. LP is characterized by the presence of white hyperkeratotic striae and skin lesions that are pruritic, purple, planar, and polygonal. It commonly affects the skin, hair, nails, and mucous membranes. The three major subtypes of LP include cutaneous LP, mucosal LP including oral LP (OLP) and genital LP, and appendageal LP [1].

OLP is the most common phenotype of LP, with an estimated prevalence between 0.5% and 2% in the general adult population [2]. OLP has two common clinical variants: erosive-atrophic (EA-OLP) and non-erosive-atrophic forms (non-EA-OLP). EA-OLP includes reticular, plaque, and papule-based forms, while non-EA-OLP includes ulcerative, atrophic, and bullous forms [2]. Importantly, OLP has a proportional risk of malignant transformation of 1.43%, most commonly progressing to oral squamous cell carcinoma [3, 4].

Diagnosis of OLP currently relies primarily on clinical examination, including identification of hallmark symptoms such as Wickham striae, erythematous changes, and painful lesions involving the buccal mucosa, tongue, gums, and lips. Histopathological confirmation is strongly recommended but is an invasive approach that requires a tissue biopsy [1]. Importantly, this diagnostic approach does not fully capture the heterogeneity among different subtypes, progression of the disease, or the underlying immunopathology of OLP. Additionally, despite its clinical significance, the etiology and pathogenesis of OLP are not completely understood. The current understanding of the disease mechanism is that CD8+ T-cells cause apoptosis of basal keratinocytes, but there are also contributions from microbial infection, trauma to the mucosa, and genetic predisposition [5]. The absence of validated, non-invasive biomarkers limits early detection and the development of targeted therapies.

Extracellular vesicles (EVs) are small, cell-derived lipid bilayer-enclosed particles, including exosomes and ectosomes [6]. They play vital roles in intracellular communication through the transfer of proteins, lipids, and nucleic acids [7]. Their molecular cargo reflects cell-state-specific biomarkers, which makes EVs compelling candidates for minimally invasive biomarker discovery in immune-mediated diseases. They have been extensively investigated as potential diagnostic biomarkers through disease-specific EV cargo signatures in biofluids, as prognostic biomarkers reflective of disease severity and progression, and as therapeutic agents through acting as delivery vehicles that can influence inflammatory and immune responses [8-10]. EVs have also been implicated in disease pathogenesis via intercellular transfer across a range of inflammatory and autoimmune conditions [11, 12].

Several studies have explored EVs as diagnostic, prognostic and functional biomarkers for OLP, while others have investigated commonly detected bacterial EVs as potential mediators of disease pathogenesis and progression. Yet a comprehensive systematic review synthesizing such evidence is not currently available. As such, we provide here a detailed systematic review & meta-analysis to evaluate EVs as biomarkers and mechanisms of disease pathogenesis and progression in LP.

## Methods

### Study design

This systematic review was conducted in accordance with the Preferred Reporting Items for Systematic Reviews and Meta-Analyses (PRISMA) guidelines. As the review used only anonymized, previously published data and involved no direct participation of human subjects or collection of personal information, institutional ethical approval was not required. The study protocol was not registered.

### Data sources and search strategy

A comprehensive literature search was conducted across PUBMED and EMBASE using search terms related to LP and its clinical subtypes, covering all results from database inception through June 27th, 2026. Two investigators (SN, ES) independently screened titles, abstracts, and full-text articles for eligibility. To ensure comprehensive review, reference lists of included studies were manually examined, and supplementary searches were conducted via Google Scholar to identify studies not indexed in the primary databases. Any discrepancies between reviewers were resolved through discussion with a third investigator (HBT). The complete search strategy can be accessed in **Table S1**.

### Eligibility criteria

A broad search strategy encompassing EVs across all LP subtypes was employed to maximize the capture of relevant literature. Studies were eligible for inclusion if they examined EVs in the context of LP, regardless of whether participants were evaluated as the primary study population or as a subgroup within a broader disease cohort.

### Data extraction and estimation

Data extraction was performed independently by multiple investigators (SN, ES), with cross-verification to ensure completeness and accuracy. A final review was conducted by an additional investigator (HBT). Extracted information included study identifiers (author, year, country, PMID), species, biomarker classification and intended clinical application (diagnostic, prognostic, predictive, prognostic, intervention or mechanistic), and evidence level. Preanalytical variables were recorded where available, including fasting status, collection tube type, platelet depletion procedures, lysis and extraction methods, and EV storage conditions. Information on sample source, EV isolation, characterization and confirmation methodology, and quantification platforms was also collected. Cohort-level variables included sample sizes of disease and control groups, demographic characteristics (age and sex), and disease severity metrics, disease duration, and disease stage. Additional variables extracted included whether studies reported quantitative biomarker data, receiver operating characteristic analyses, correlation analyses, predictive or prognostic modeling, and differential expression data for miRNAs, EV-associated proteins, cytokines, and surface markers.

Weighted means were calculated for mean age and female percentage across all studies. Only studies that reported both the demographic variable of interest and the corresponding group-specific sample size were included in each analysis. For each subgroup, the total sample size was first calculated by summing all eligible participants across studies reporting complete data for the variable of interest. For mean age, each study was given a weight, calculated by dividing the number of participants in each group by the total sample size for that subgroup. These weights were then multiplied by the corresponding mean age reported in each study. The resulting values were summed to give the overall weighted mean for each subgroup. For female percentage, the total number of eligible females was summed within each subgroup. The reported percentage of females in each study was multiplied by the corresponding study sample size. These values were summed across studies and divided by the total sample size.

One study separated individuals with OLP/Oral Lichenoid Lesions (OLL) and controls into 9 total OLP/OLL cohorts and 6 control cohorts for analysis. Weighted means were calculated for each cohort and then combined to obtain overall study values used in the pooled analyses.

From two studies, we reconstructed individual-level data from published figures using WebPlotDigitizer, which was used to perform in-house receiver operating characteristic (ROC) analyses via binary logistic regression. From each figure, individual data points were manually identified and plotted over the original scatter plot within WebPlotDigitizer, generating a set of x- and y-coordinate values. Using these values, ROC analyses were performed to obtain the area under the curve (AUC), optimal cut-off value, sensitivity, specificity, 95% confidence interval (CI), and p-value. The optimal cut-off was defined as the value maximizing Youden’s index.

Byun et al. did not report ROC analyses, so individual-level values for miR-4484 from their Figure 3A were digitized and used to generate ROC curves. As this figure contained a discontinuous y-axis with a scale break between 10 and 40, data were reconstructed using separate axis calibrations across three ranges (0-10, 10-40, and 40-100). Yao et al. reported AUCs for *GJA1* and Cx43 from ROC curves presented in their Figures 3B and 3D, respectively, but did not provide cut-off values, sensitivities, specificities, CIs, or a combined biomarker model. Individual-level values for both biomarkers were digitized from the scatter plots presented in their Figures 3A (*GJA1*) and 3C (Cx43). The validity of the reconstructed data set was confirmed by the close agreement between our calculated AUCs (*GJA1:* 0.741; Cx43: 0.785) and those reported by the authors in Figures 3B and 3D (*GJA:* 0.74; Cx43: 0.79).

### Risk of bias assessment

Risk of bias was evaluated using a modified version of the Newcastle-Ottawa Scale adapted for cross-sectional studies. Cross-sectional studies that compared EV biomarkers between patients with LP and controls or individuals with other disease subtypes were classified as diagnostic studies. This categorization reflected the intended application of EV biomarkers rather than the primary design or objectives of the parent study. Risk of bias assessments were independently performed by SN and ES with final verification by HBT. No discrepancies were identified among reviewers.

### Statistical analysis

The two most common models for meta-analyses of diagnostic accuracy are the hierarchical summary ROC (HSROC) model and the bivariate random effects meta-analysis model (BRMA) [16], both of which are equivalent when no covariates are included [17]. We used the BRMA model if more than 3 studies were available and a univariate fixed-effects model if 3 or fewer studies were available [18]. We fit the data with the unstructured or structured covariance matrix based on the lowest combination of Akaike information criteria (AIC) and Bayesian information criteria (BIC). We selected the model with the lowest AIC in cases of a tie. In the resulting HSROC curve, proximity of the SROC line and mean point to the upper left quadrant indicates higher diagnostic accuracy.

Heterogeneity was evaluated using the *I*^2^ statistic as described by Zhou & Dendukuri [19]. Publication bias [20] was assessed using Begg’s rank correlation, Egger’s and Deeks’ regression, and the trim-and-fill method [21]. Studies reporting a sensitivity and/or specificity of 1.00 were excluded from the publication bias assessment. Meta-regression analyses using included categorical variables were additionally performed to examine the potential impact of different covariates on the absolute sensitivity and specificity.

The BRMA model and associated meta-regressions were conducted using the *metadta* command in Stata [18], which has been extensively validated against alternative packages [22]. All remaining analyses were conducted in RStudio (version 2022.12.0 □+ □353).

## Results

### Cohort characteristics

A total of 10 studies [13-15, 23-29] were identified as meeting inclusion criteria and included in the systematic review (**Figure 1**). Detailed information on each study is provided in **Table 1**. Six studies evaluated EV biomarkers in a diagnostic context, while two included prognostic analyses. No studies evaluated EV-associated biomarkers in response to a treatment. Three studies included functional experiments, in which they isolated EVs from individuals with OLP and administered them into Jurkat cells and oral keratinocytes pretreated with LPS. Two studies were mechanistic studies, in which bacteria-derived EVs were applied to *in vitro* models to assess downstream biological effects and inflammatory signaling responses compared to those found in OLP lesions.

**Figure 1.**
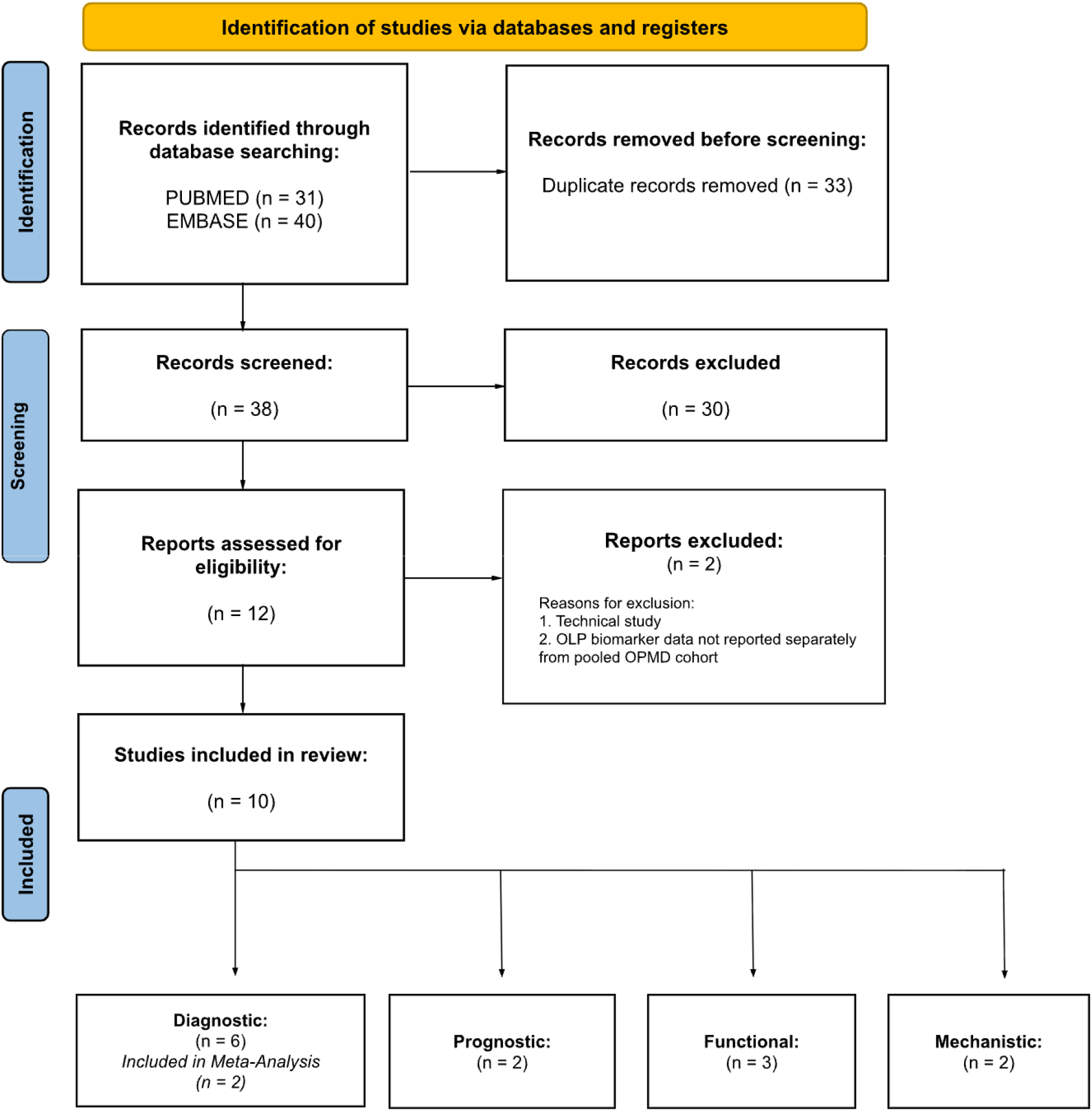
PRISMA flow diagram.

Two studies were excluded after the full-text screen. Peng et al. was excluded as it was a technical comparative study and did not quantify OLP-associated EV biomarkers. Yap et al. evaluated a panel of select miRNAs from individuals with oral squamous cell carcinoma and oral potentially malignant disorders, but it was excluded because no biomarker data were reported for participants with OLP as an individual cohort.

The ten studies included a total of 298 individuals with LP (weighted mean age: 50.7 years; 61.5% female), 194 controls (weighted mean age: 47.8 years; 58.5% female). Individuals with LP were entirely of the OLP subtype except for one study which grouped individuals with OLP/OLL together. With the exclusion of that study, there were a total of 261 individuals with OLP (weighted mean age: 50.7; 61.4% female).

The eight clinical biomarker studies included a total of 283 individuals with LP (weighted mean age: 50.7 years; 61.0% female), 179 controls (weighted mean age: 49.8 years; 57.9% female). Individuals with LP were entirely of the OLP subtype with the exception of one study which grouped individuals with OLP/OLL together. With the exclusion of that study, there were a total of 246 individuals with OLP (weighted mean age: 50.7; 60.9% female).

The two mechanistic studies included a total of 15 individuals with LP (weighted mean age: 51.0 years), all of whom were individuals with OLP, and 15 controls. For the mechanistic experiments in Wang et al., primary fibroblasts were isolated from a subset of 8 patients with OLP and 8 healthy controls rather than the full clinical cohort. These sample sizes were used for weighting in the pooled analyses. Kim et al. did not report the age of the control group or the sex distribution of either cohort.

Demographic information including BMI, other diseases, surgeries, disease stage, and disease duration were not provided in any of the 10 studies. One study reported symptoms/comorbidities of each cohort: 16/68 of OLP and 9/57 of controls had digestive diseases, and 55/68 of OLP and 42/57 of controls had dental calculi. Additionally, six of the 10 studies specified that participants were treatment-naive within a specified number of months (ranging from one to six) before enrollment. Three studies also discussed inclusion of severity scores to assess individuals with OLP, but none provided a breakdown of where patients ranked within the severity scale used. All studies clinically diagnosed and histopathologically confirmed OLP according to the modified WHO diagnostic criteria [32].

### EV characteristics

In the eight studies investigating EVs as biomarkers (**Table 1**), EVs were isolated from various sources, including plasma (n=2 studies), saliva (n=2 studies), oral tissue (n=1 study), primary T-culture supernatant derived from peripheral blood mononuclear cells (PBMCs) (n=2 studies), and serum (n=1 study). EV isolation methods varied across studies and included UC (n=3), EV isolation kits such as ExoQuick and exoEasy Maxi Kit (n=3), size exclusion chromatography (SEC) followed by ultrafiltration (UF) (n=1), and EV Enrichment Kit followed by immunoprecipitation of CD63+ EVs (n=1). EV characterization techniques included flow cytometry (FC) (n = 3), nanoparticle tracking analysis (NTA) (n = 7), transmission electron microscopy (n = 8), and Western blots (WB) (n = 5). FC panels primarily assessed EV surface markers such as CD9 and CD63, while WB commonly evaluated EV-enriched proteins, such as ALIX, CD9, CD63, and TSG101, along with negative controls such as GAPDH, β-actin, or calnexin. EV biomarker quantification methods included FC (n=3), ELISA (n=1), WB (n=1), immunohistochemistry assessing oral lesion tissue expression of the EV marker CD63 (n=1), multiplexed microarray analysis (n=1), Luminex xMAP-based cytokine assays (n=1), miRNA sequencing (n=2), and qPCR (n=3). Two studies additionally performed proteomic profiling using label-free LC-MS/MS.

In the mechanistic studies (**Table 1**), EVs were isolated from bacterial culture supernatants. The EV isolation method was UC followed by 0.22-μm filtration, UF, and either usage of an ExoBacteria OMV Isolation Kit or a final UC step. EV characterization techniques included NTA (n=2), high-performance liquid chromatography-mass spectrometry (HPLC-MS) (n=1), and scanning electron microscopy (n=1).

### Risk of bias

All included studies were cross-sectional in design, and methodological quality was assessed using a modified Newcastle-Ottawa Scale. Overall, studies demonstrated fair methodological quality, with recurrent limitations related to sample representativeness, selection of control samples, control for confounding variables, and pre-analytical handling. Among the included studies (**Table S2**), 6 studies had reasonably representative samples of individuals with LP across varying levels of disease severity and activity, while 5 studies selected controls from the same population as participants with LP. All studies used adequate methods for ascertainment of exposure, with OLP diagnoses established using the modified WHO criteria [32]. However, adjustment for variables such as age, sex, BMI, or disease severity was generally absent or inadequately reported. All studies also utilized targeted analytical methods, including qPCR or flow cytometry, to measure EV biomarkers, and 7 studies confirmed EV isolation using at least two characterization techniques. Only one study adequately described pre-analytical EV handling procedures, including factors such as fasting status, collection timing, collection tubes, platelet depletion, freezing conditions, or lysis methods. Overall, these findings indicate moderate methodological quality, although heterogeneity in EV isolation, processing, characterization, and confounder control may limit the reproducibility and interpretation of results.

#### Diagnostic

##### RNA

Three studies evaluated EV-associated RNA biomarkers in a diagnostic context (**Table 2**). Two studies performed bulk sequencing-based approaches followed by qPCR validation of selected candidates. They found a total of 11 miRNAs to be upregulated and 10 to be downregulated. However, there was no overlap of differentially expressed miRNAs across studies, highlighting substantial heterogeneity in reported findings **(Table 2)**.

Byun et al. performed miRNA microarray analysis, identifying 8 upregulated and 8 downregulated miRNAs in OLP compared with control EVs. They attempted to validate these results by performing qPCR on 3 candidate biomarkers: miR-1246, miR-1290 and miR-4484. miR-4484 was significantly upregulated in OLP EVs compared with controls, while miR-1246 and miR-1290 showed no significant changes. Meanwhile, Peng et al. used miRNA microarrays and identified 3 upregulated and 2 downregulated miRNAs in OLP compared with control EVs. They attempted to validate all five candidates by qPCR. Both miR-34a-5p and miR-130b-3p were significantly upregulated, and miR-301b-5p was significantly downregulated in EVs from individuals with OLP compared with controls. miR-29c-3p and miR-144-3p showed no significant differences. Yao et al. on the other hand, used only qPCR to compare *GJA1*, which encodes Connexin 43 (Cx43), in reticular, hyperemia, and erosion type OLP with control EVs. They found *GJA1* to be upregulated in all three OLP subtypes compared with controls and between the erosion and hyperemia type OLP. Overall, the absence of overlapping miRNAs and mRNAs across studies limits conclusions about specific EV-associated RNAs, but several candidate targets (e.g., *GJA1*, miR-4484) warrant further investigation as potential diagnostic biomarkers for OLP.

##### Protein

Two studies employed proteomic approaches to identify differentially expressed EV proteins in OLP compared with controls (**Table 3**). A total of 133 proteins were upregulated, and 204 proteins were downregulated. However, one study reported only aggregate numbers of differentially expressed proteins and did not provide a complete list of individual protein identities, limiting the ability to assess overlap between the studies. Among the proteins reported in both studies, no overlap in the upregulated or downregulated proteins was observed.

Sun et al. first compared proteomic profiles of oral tissue-derived EVs isolated from individuals with OLP/OLL (n=29 tissues pooled into 6 samples) and controls (n=18 tissues pooled into 3 samples) using label-free quantitative nano-LC-MS/MS. They identified a total of 141 proteins, of which 10 were differentially expressed (fold change >2 or <0.5, adjusted p<0.05). Of the differentially expressed proteins, five were upregulated in OLP EVs, including elongation factor 1-alpha 1 (EIF1A), fibrinogen alpha chain (FGA), lamin isoform A (LMNA), protein disulfide isomerase family A member 3 (PDIA3), and 40S ribosomal protein (RPS40). In contrast, five proteins were downregulated, including collagen alpha-3 chain (COL3A1), guanine nucleotide-binding protein subunit gamma-12 (GNG12), katanin p80 repeat-containing subunit B1 (KATNB1), mitochondrial 2-oxoglutarate/malate carrier protein (SLC25A11), and plakophilin-1 (PKP1). PDIA3 was selected for further validation due to its known involvement in antigen processing and presentation and enrichment in several immune-related pathways. WB subsequently confirmed the upregulation of PDIA3 in oral tissue-derived EVs isolated from a validation cohort composed of 8 individuals with OLP/OLL pooled into 3 groups (n=4, 2, and 2) compared with6 controls pooled into 3 groups (n=2, 3, and 1).

Similarly, Wang et al. performed label-free quantitative LC-MS/MS proteomics on serum-derived EVs isolated from 10 individuals with erosive OLP, 10 individuals with non-erosive OLP, and 10 controls. A total of 327 proteins met the criteria for differential expression (fold change of ≥ 1.2 or ≤ 0.8333 and adjusted p<0.05). Compared with controls, they indicate that 138 differentially expressed proteins were identified in the overall OLP group, including 39 upregulated and 99 downregulated proteins (**Table 3**). Subgroup analyses identified 70 differentially expressed proteins in non-erosive OLP compared with control EVs (30 upregulated, 40 downregulated) and 99 differentially expressed proteins in erosive OLP compared with controls (52 upregulated, 47 downregulated). Additionally, 20 differentially expressed proteins (13 upregulated and 7 downregulated) distinguished erosive from non-erosive OLP. However, only a subset of these differentially expressed proteins was reported, and individual protein-level data were not provided.

Beyond proteomic profiling, two studies quantified specific EV-associated biomarkers using targeted methods. Yang et al. measured the expression of 23 cytokines in T cell-derived EVs from individuals with OLP (n=24) and controls (n=19) using a Luminex xMAP-based assay. They discovered lower levels of IL-1β, IL-5, and IFN-γ and higher levels of IL-7, IL-10, IL-12, and especially IL-17 in OLP compared with control EVs. In a separate study, Yao et al. quantified Connexin 43 (Cx43), the protein product of *GJA1*, in salivary EVs isolated from individuals with OLP (n=68) and controls (n=57) using an ELISA. They showed that Cx43 levels were upregulated in OLP compared with control EVs, as well as in reticular (n=27), hyperemia (n=21), and erosion (n=20) type OLP compared with control EVs.

##### Diagnostic accuracy meta-analysis

Of the included studies, only two presented figures with individual-level data points comparing biomarker levels between individuals with OLP and controls. This allowed us to reconstruct individual data points from these published figures using WebPlotDigitizer (**Figure S1 and S2**) to perform in-house ROC analyses using a binomial logistic regression and obtain the sensitivity and specificity at the cutoff that optimizes Youden’s index (see Methods for further details). Using reconstructed data from Byun et al., miR-4484 yielded an AUC of 0.81 (95% CI = 0.61-1.00, p = 0.14), with a sensitivity of 71% and specificity of 100%. Although Yao et al. reported ROC analyses for *GJA1* (AUC = 0.74) and Cx43 (AUC = 0.79), they did not provide the accompanying cut-off values, sensitivities, specificities, or confidence intervals. As such, we reconstructed the data (Figure S2) and performed ROC analyses. In our hands, *GJA1* yielded an AUC of 0.741 (95% CI = 0.656-0.826, p < 0.0001), with a sensitivity of 48.5% and specificity of 87.7%. Cx43 yielded an AUC of 0.785 (95% CI = 0.707-0.86, p < 0.0001), with a sensitivity of 70.6% and specificity of 77.2%. A combined biomarker model incorporating both *GCJA1* and Cx43 yielded an AUC of 0.892 (95% CI = 0.835-0.949, p < 0.0001) with a sensitivity of 79.4% and specificity of 89.5%.

To identify the best diagnostic biomarker(s) for distinguishing OLP from controls, we pooled these statistics using a diagnostic accuracy meta-analysis using a BRMA model (**Figure 2**). The overall pooled AUC was 0.89 with a sensitivity of 78% (95% CI: 0.78-0.96) and specificity of 90% (95% CI: 0.80-0.96). However, because only two studies were included in this analysis, these results remain preliminary with limited interpretation, and heterogeneity could not be assessed.

**Figure 2.**
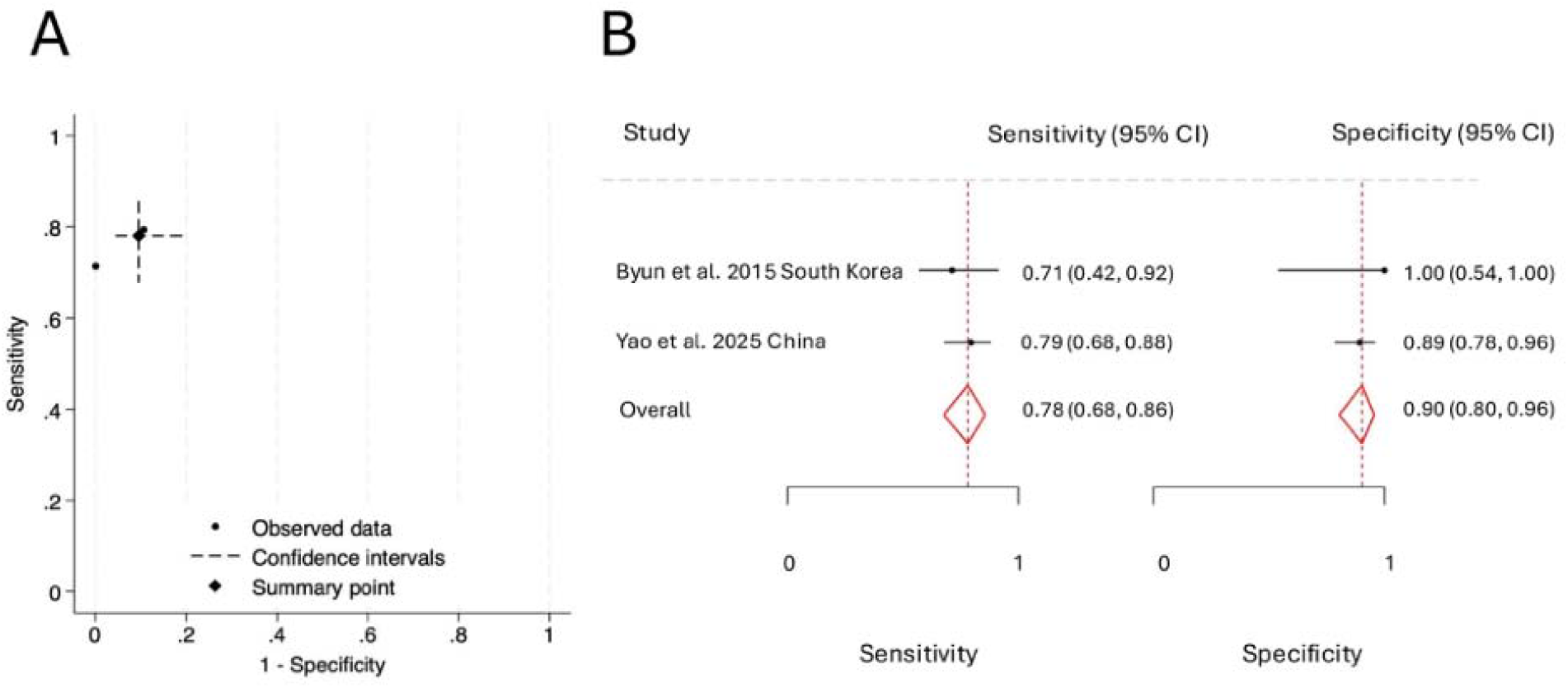
Diagnostic accuracy of extracellular vesicle (EV)-derived biomarkers for distinguishing OLP. (A-B) Diagnostic performance of EV biomarkers for distinguishing OLP from controls. (A) Summary receiver operating characteristic (SROC) plot showing individual study estimates, confidence intervals, and the pooled summary point. (B) Forest plots of sensitivity and specificity for each included study, with point estimates and 95% confidence intervals shown for sensitivity (left panel) and specificity (right panel); red diamonds and red dashed lines indicate pooled summary estimates.

##### Prognostic

Two studies investigated EV-associated biomarkers in relation to disease severity, suggesting their potential as predictors of OLP activity. Peng et al. examined the relationship between EV-miRNAs and Reticular, Atrophic, Erosive (RAE) scores, which reflect the severity of OLP. miR-34a-5p showed a moderate positive correlation with disease severity (r = 0.6093, P = 0.012), whereas miR-130b-3p (r = −0.278, P = 0.31) and miR-301b-3p (r = −0.2986, P = 0.26) showed weak, non-significant negative correlations (**Table 2**). Similarly, Yao et al. performed correlation analyses and found that *GJA1* and Cx43 were positively associated with disease activity scores (DAS) (*GJA1*: *r* = 0.51, P < 0.001; Cx43: r = 0.57, P < 0.00) (**Table 2 and 3)**. They also found that *GJA1* was positively correlated with IL-17 (r = 0.35, P < 0.001) and TNF-α (r = 0.32, P < 0.001) (**Table 2**). Cx43 was also positively correlated with IL-17 (r = 0.42, P < 0.001) and TNF-α (r = 0.37, P < 0.001) (**Table 3)**. This suggests that *GJA1* and Cx43 may be indicators of underlying inflammation in OLP.

##### Functional

Three articles examined EVs as functional biomarkers (**Table 4**). Peng et al. isolated plasma-derived EVs from individuals with OLP (n=12) and controls (n=6) and incubated them with Jurkat T-cells for 12, 24, and 48 hours. They demonstrated that OLP-derived EVs increased T-cell proliferation (CCK-8 assay), reduced T-cell apoptosis (Annexin V assay), enhanced T-cell migration (Transwell assay), and modified T-cell cytokine polarization by increasing IFN-γ and decreasing IL-4 secretion (ELISA). These findings suggest that circulating OLP EVs may actively push T cells towards a higher inflammatory state, contributing to disease progression. Yang et al. isolated T cell-derived EVs from individuals with OLP (n=87) and controls (n=44) and incubated them with Jurkat T-cells for 48 hours. OLP T-cell-derived EVs increased the production of MIP□1α/β, along with IL-6, IL-10, and IL-17A (Luminex assay) when compared with control EVs, indicating a pro-inflammatory EV-mediated response. In another study, Yang et al. isolated T-cell-derived EVs from individuals with OLP (n=24) and controls (n=19) and cocultured them separately with human oral keratinocytes pretreated with LPS for 24 hours and Jurkat T-cells for 48 hours. They conducted two functional assays on the keratinocytes, concluding that OLP and control T-cell-derived EVs induced keratinocyte apoptosis (Annexin V assay) compared with PBS controls, but OLP-derived EVs did not significantly increase apoptosis compared with controls. Additionally, they showed that T-cell-derived EVs reduced cell numbers and led to morphological changes in keratinocytes, including cell shrinkage and irregular borders. On the other hand, the proliferation of treated Jurkat cells (CCK-8 assay) was relatively increased in the OLP EV-treated group compared with the control EV-treated group, and the OLP EV-treated group did not significantly change Jurkat T-cell proliferation compared with PBS-treated controls.

##### Mechanistic

Two studies investigated EVs as potential mechanistic mediators of OLP disease pathogenesis and progression (**Table 5**).

Kim et al. isolated EVs from key periodontal pathogens involved in OLP pathogenesis, *Porphyromonas gingivalis* (*P. gingivalis*) and *Aggregatibacter actinomycetemcomitans* (*A. actinomycetemcomitans*). HOK-16B human oral keratinocytes were incubated with PBS or bacterial EVs for 24 hours. They showed that bacterial EVs derived from *A. actinomycetemcomitans* and *P. gingivalis* increased expression of the pro-inflammatory cytokines *IL-1*β, *IL-6, IL-18* and *TNF-*α (qPCR), as well as increased STAT3 phosphorylation and NLRP3 activation (WB) in treated keratinocytes. These findings corresponded directly with observations in OLP lesions, where *IL-1*β, *IL-6, IL-8* and *TNF-*α (qPCR) and phosphorylated STAT3 levels (IF) were elevated in the epithelium compared with non-OLP tissues. These findings suggest that bacterial EVs may contribute to OLP pathogenesis by activating the inflammatory STAT3 pathway.

Similarly, Ji et al. identified increased *Parvimonas micra* (*P. micra*) in OLP buccal mucosa compared with controls. As such, they isolated EVs from *P. micra* and incubated them with healthy human oral fibroblasts to assess functional effects. They showed that *P. micra*-derived EVs increased TNF-α expression (ELISA), activated NF-κB signaling (WB), and disrupted autophagic flux, as demonstrated by increased LC3B-II and SQSTM1 expression together with accumulation of autophagosomes and reduced autolysosome formation (mCherry-GFP-LC3 assay). Proteomic analysis (HPLC-MS) identified DnaK as a key EV cargo protein. These results were consistent with those observed in OLP-derived fibroblasts, which showed activation of inflammatory pathways such as NF-κB, increased TNF-α expression, and impaired autophagic flux compared with control fibroblasts. These findings support a potential role for *P. micra*-derived EVs in OLP pathogenesis by promoting inflammation and disrupting autophagy in oral fibroblasts.

## Discussion

Early and accurate diagnosis of LP poses a significant challenge due to the lack of validated non-invasive biomarkers and an incomplete understanding of its pathogenesis. EVs have emerged as promising candidates to address these challenges due to their ability to carry active molecular cargo and potential contributions to disease pathogenesis [7]. This systematic review and meta-analysis provides a comprehensive synthesis of EV-associated biomarkers and their functional and mechanistic roles in OLP using 10 studies [13-15, 23-29]. EV-associated miRNAs and proteins have potential as diagnostic and prognostic biomarkers, and functional and mechanistic studies contribute to an understanding of how EVs may actively drive inflammatory pathways in OLP. Several candidate biomarkers, including miR-4484, *GJA1*, and Cx43 (**Tables 2-3**), through ROC analyses, demonstrated good diagnostic utility, and miR-34a-5p, *GJA*, and Cx43 showed a significant positive correlation with severity of OLP. Additionally, the included studies show that EVs isolated from individuals with OLP carry functional biomarkers that can alter the behavior of healthy recipient cells by promoting inflammation, immune activation, cell proliferation, migration, and resistance to apoptosis (**Table 4**). EVs released by bacterial strains commonly found in OLP lesions also caused inflammatory responses, suggesting that these EVs may also contribute to disease pathogenesis and progression (**Table 5**).

EV-associated miRNAs (**Table 2**) demonstrated significant heterogeneity, with no differentially expressed miRNAs overlapping across studies. However, several candidates showed preliminary promise, including miR-4484, as well as miR-34a-5p and miR-130b-3p, which were validated by qPCR after initial microarray identification. miR-4484 also demonstrated good diagnostic utility for distinguishing individuals with OLP from controls (AUC = 0.81). Although EV-based findings were not replicated across studies, these miRNAs have been independently reported in other related inflammatory diseases, including systemic sclerosis, where miR-4484 has been identified as a promising circulating biomarker and linked to fibrotic pathways such as TGF-β signaling. The lack of reproducibility across studies is likely attributed to differences among studies in regard to sample source, EV isolation method and characterization, and cohort composition rather than a true absence of biologically relevant EV-associated biomarkers.

EV-associated protein findings (**Table 3**) concentrate on several overlapping pathological themes, including antigen presentation, immune activation, and inflammatory signaling. PDIA, which is involved in antigen processing and presentation, was identified as the most immunologically relevant upregulated protein amongst oral tissue-derived OLP EVs. This suggests its potential role in driving immune activation within OLP lesions. Additionally, EV-associated Cx43, a gap junction protein involved in inflammatory intercellular communication, was positively correlated with both OLP disease severity and the pro-inflammatory cytokines IL-17 and TNF-α. These results are biologically relevant, as Cx43 has been implicated in T-cell activation, proliferation, and cytokine signaling, with its inhibition leading to reduced cytokine secretion. Together, these findings suggest that EV-derived Cx43 may contribute to the transmission of inflammatory signals between oral mucosal cells. In our analysis, the combination of *GJA1* (the mRNA encoding Cx43) and Cx43 yielded the strongest diagnostic performance (AUC = 0.892), further suggesting that it is a clinically relevant biomarker.

Functional studies suggest that EVs may actively contribute to inflammation in OLP rather than only reflecting disease activity (**Table 5**). Plasma-derived OLP EVs have been shown to increase T cell proliferation and migration, reduce apoptosis, and promote a pro-inflammatory cytokine profile.

Meanwhile, T-cell-derived EVs have been shown to increase production of MIP □1α/β and cause keratinocyte apoptosis without direct cell contact. Together, these findings suggest that EVs may contribute to immune cell recruitment and epithelial injury in OLP. In addition, findings from mechanistic studies demonstrate that bacterial EVs further amplify inflammatory signalling. EVs from periodontal bacteria such as *P. gingivalis* and *A. actinomycetemcomitans* promote expression of pro-inflammatory cytokines such as *IL-1*β, *IL-6, IL-18* and *TNF-*α and activate STAT3 and NLRP3 inflammatory pathways in oral keratinocytes, while *P. micra*-derived EVs separately activated NF-κB and suppressed autophagy in fibroblasts, contributing to sustained inflammatory responses. Overall, results from both functional and mechanistic studies support a model in which both host- and bacteria-derived EVs work together to amplify immune activation and tissue damage in OLP.

The findings of this review align with findings of several recent studies that demonstrate the relevance of EVs as biomarkers and therapeutic agents in dermatological disorders. Studies have identified disease-associated EV miRNAs and proteins in chronic inflammatory skin diseases such as psoriasis, atopic dermatitis, and psoriatic arthritis. For instance, IL-17A-positive EVs have been associated with psoriasis severity, while serum microbial EVs have shown potential as diagnostic biomarkers in atopic dermatitis. EVs are also currently being investigated as therapeutic agents for inflammatory skin conditions, such as in atopic dermatitis where adipose-derived mesenchymal stem cell EVs have been shown to reduce disease severity and reduce inflammatory cytokine expression. Collectively, these findings support EVs as a promising platform for both biomarker discovery and therapeutic tools in inflammatory dermatological diseases like LP.

This review has several limitations. With only 10 included studies, the sample size was small, and most candidate biomarkers were only reported in one study, preventing cross-study comparisons. There was also significant methodological heterogeneity in EV isolation, characterization, sample sources, and biomarker quantification. No studies evaluated EV biomarkers in relation to treatment or longitudinal outcomes, which limits conclusions about predictive ability. While all of the studies included individuals with OLP as their study population, one study grouped individuals with OLP and OLL together without providing individual data for each subgroup, which may have introduced heterogeneity into the pooled data. Due to the availability of current literature, although our goal was to investigate biomarkers for LP and all its subtypes, we did not find any studies investigating EVs as biomarkers for LP or other subtypes other than OLP. Additionally, only two studies were included in the diagnostic accuracy meta-analysis, both of which were based on data estimated from study figures replicated using WebPlotDigitizer, limiting the utility and precision of these results. Future studies should prioritize standardized EV methodologies, biomarker validation in larger and more homogeneous cohorts, and longitudinal designs to better characterize treatment outcomes and disease progression. Authors should also be encouraged to make their datasets publicly available or provide high-resolution figures with overlaid individual data points to allow other researchers to independently reconstruct analyses, validate reported findings, and perform pooled analyses across studies. Overall, current evidence supports EVs as potential biomarkers and therapeutic candidates in LP.

## Conclusion

This systematic review and meta-analysis provide the first comprehensive synthesis of EV-associated biomarkers and mediators of disease pathogenesis and progression in LP, with findings concentrated in OLP. Overall, EV-associated miRNAs and proteins appear to be promising biomarker candidates in LP, reflecting key inflammatory and immune-related processes underlying disease pathophysiology. Future studies should prioritize larger longitudinal cohorts and standardized methodologies to validate candidate EV biomarkers and better determine their clinical utility. These studies will be important to assess their predictive value in patients, while clarifying whether EV-based approaches can improve diagnostic precision, disease monitoring, and development of targeted therapies in LP.

## Data Availability

Data analyzed are included in the article.

## Acknowledgement

None

