## Supplementary material for "Extracellular vesicles as biomarkers and disease mediators in lichen planus: a systematic review & meta-analysis": Table S1

**Table S1: Complete Search strategy using PUBMED and EMBASE. The search was performed from the date of inception until June 27^th^, 2026.**

| PUBMED | ( "extracellular vesicle*"[Title/Abstract] OR exosome*[Title/Abstract] OR ectosome*[Title/Abstract] OR microvesicle*[Title/Abstract] OR "apoptotic bod*"[Title/Abstract] OR microparticle*[Title/Abstract] ) AND ( "Lichen Planus"[Mesh] OR "lichen planus"[Title/Abstract] OR "oral lichen planus"[Title/Abstract] OR "cutaneous lichen planus"[Title/Abstract] OR "erosive lichen planus"[Title/Abstract] OR "hypertrophic lichen planus"[Title/Abstract] OR "lichen planopilaris"[Title/Abstract] ) NOT ( review[Publication Type] OR systematic review[Title/Abstract] OR meta-analysis[Publication Type] ) |
| --- | --- |
| EMBASE | ('extracellular vesicle*':ti,ab OR exosome*:ti,ab OR ectosome*:ti,ab OR microvesicle*:ti,ab OR 'apoptotic bod*':ti,ab OR microparticle*:ti,ab) AND ('lichen planus'/exp OR 'lichen planus':ti,ab OR 'oral lichen planus':ti,ab OR 'cutaneous lichen planus':ti,ab OR 'erosive lichen planus':ti,ab OR 'hypertrophic lichen planus':ti,ab OR 'lichen planopilaris':ti,ab) NOT ('review'/it OR 'systematic review':ti,ab OR 'meta analysis'/it) |

**Table S2. Quality assessment of cross-sectional studies.** Methodological quality of included cross-sectional studies was evaluated using a modified Newcastle–Ottawa Scale (NOS) across domains of sample representativeness, selection of control sample, ascertainment of exposure, control for sex or age, control for additional confounding variables, analysis of biomarkers, confirmation of EVs and pre-analytical sample handling. Each domain was rated as high quality (green), unclear or partially reported (yellow), or poor quality (red). **D1** – sample representativeness; **D2** – selection of control sample; **D3** – ascertainment of exposure; **D4** – control for sex or age; **D5** – control for additional confounding variables; **D6** – analysis of biomarkers; **D7** – confirmation of EVs; **D8** – pre-analytical sample handling.

|  | **D1** | **D2** | **D3** | **D4** | **D5** | **D6** | **D7** | **D8** |
| --- | --- | --- | --- | --- | --- | --- | --- | --- |
| **Byun et al. 2015 Korea** | **-** | **+** | **+** | **+** | **?** | **+** | **+** | **-** |
| **Peng et al. 2019a China** | **+** | **+** | **+** | **+** | **?** | **+** | **+** | **-** |
| **Peng et al. 2019b China** | **+** | **-** | **+** | **+** | **?** | **+** | **+** | **-** |
| **Yang et al. 2020 China** | **+** | **-** | **+** | **+** | **?** | **+** | **+** | **-** |
| **Yang et al. 2021 China** | **+** | **+** | **+** | **+** | **?** | **+** | **-** | **-** |
| **Sun et al. 2021 China** | **+** | **+** | **+** | **+** | **?** | **+** | **+** | **--** |
| **Yao et al. 2025 China** | **+** | **+** | **+** | **+** | **+** | **+** | **+** | **-** |
| **Wang et al. 2025 China** | **-** | **-** | **+** | **?** | **?** | **+** | **+** | **+** |
